# Conceptualising the Episodic Nature of Disability among Adults Living with Long COVID: A Qualitative Study

**DOI:** 10.1101/2022.11.12.22282116

**Authors:** Kelly K. O’Brien, Darren A. Brown, Kiera McDuff, Natalie St. Clair-Sullivan, Patricia Solomon, Soo Chan Carusone, Lisa McCorkell, Hannah Wei, Susie Goulding, Margaret O’Hara, Catherine Thomson, Niamh Roche, Ruth Stokes, Jaime H. Vera, Kristine M. Erlandson, Colm Bergin, Larry Robinson, Angela M. Cheung, Brittany Torres, Lisa Avery, Ciaran Bannan, Richard Harding

## Abstract

**Objectives:** To describe episodic nature of disability among adults living with Long COVID.

**Methods:** We conducted a community-engaged qualitative descriptive study involving online semi-structured interviews and participant visual illustrations. We recruited participants via collaborator community organizations in Canada, Ireland, United Kingdom, and United States.

**Participants:** Adults who self-identified as living with Long COVID. We purposively recruited for diversity in age, gender, race/ethnicity, sexual orientation, and duration since initial COVID-19 infection.

**Main Outcome Measure(s):** We used a semi-structured interview guide to explore experiences of disability living with Long COVID, specifically health-related challenges and how they were experienced over time. We asked participants to draw their health trajectory and conducted a group-based content analysis.

**Results:** Among the 40 participants, the median age was 39 years (interquartile range: 32, 49); majority were women (63%), white (73%), heterosexual (75%), and living with Long COVID for ≥1 year (83%). Participants described their disability experiences as episodic in nature, characterized by fluctuations in presence and severity of health-related challenges (disability) that may occur both within a day and over the long-term living with Long COVID. They described living with ‘ups and downs’, ‘flare-ups’, and ‘peaks’ followed by ‘crashes’, ‘troughs’, and ‘valleys’, likened to a ‘yo-yo’ ‘rolling hills’, and ‘rollercoaster ride’ with ‘relapsing/remitting’, ‘waxing/waning’, ‘fluctuations’ in health. Drawn illustrations demonstrated variety of trajectories across health dimensions, some more episodic than others. Uncertainty intersected with the episodic nature of disability, characterized as unpredictability of episodes, their length, severity and triggers, and process of long-term trajectory, which had implications on broader health.

**Conclusions:** Among this sample of adults living with Long COVID, experiences of disability were described as episodic, characterized by fluctuating health challenges, which may be unpredictable in nature. Results help to better understand experiences of disability among adults living with Long COVID to inform healthcare and rehabilitation.

**KEY MESSAGES:**

- **What is already known on this topic:** Globally, a growing number of individuals are living with persistent and prolonged signs and symptoms following infection consistent with COVID-19, referred to as Long COVID, Post COVID-19 Condition (PCC) or Post-acute sequelae of SARS-CoV2 (PASC). Individuals living with Long COVID are experiencing a range of symptoms and impairments that impact their ability to carry out day to day activities or engage in social and community life roles.
- **What this study adds:** Disability living with Long COVID was described as episodic, characterized by fluctuations in presence and severity of health related challenges, which may be unpredictable in nature, occurring both within the day, and over the long-term of months and years living with Long COVID.
- **How this study might affect research, practice or policy:** Results will help researchers, healthcare providers, policymakers, employers, and community members to better understand experiences of disability among adults living with Long COVID, to inform future disability measurement, health and rehabilitation care and service delivery, programs and policies for insurance, return to work, and workplace accommodations.

## INTRODUCTION

More individuals are living with persistent and prolonged signs and symptoms following infection consistent with COVID-19, referred to as Long COVID or Post COVID-19 Condition (PCC) or Post-acute sequelae of SARS-CoV2 (PASC).^1 2^ According to the World Health Organization definition, Long COVID occurs usually 3 months from the onset of probable or confirmed COVID-19 with symptoms that last for at least 2 months and cannot be explained by an alternative diagnosis.^3^

An estimated 144 million individuals are living with Long COVID globally although estimates vary considerably across studies.^4-7^ A systematic review involving 50 studies concluded the pooled prevalence of Long COVID globally was 43% among hospitalized (53%) and non-hospitalized patients (34%).^2^ Systematic review evidence examining symptoms among individuals after COVID-19 reported that 38-72% were living with >1 symptom for at least 2 months from COVID-19 onset and up to 54% were living with persistent symptoms for 6 or more months.^8-12^

Long COVID is characterized by a multitude of health symptoms that affect daily functioning and social participation.^13 14^ Among the estimated 2.3 million people living with Long COVID in the UK, 72% reported their symptoms negatively impacted their daily activities.^6^ Fatigue was the most commonly reported symptom (69%), followed by difficulty concentrating (45%), shortness of breath (42%), and muscle aches (40%). A community-led online survey across 56 countries identified participants experienced an average of 56 symptoms spanning nine organ systems including fatigue, post exertional malaise (PEM) or post exertional symptom exacerbation (PESE), and cognitive dysfunction, with 86% of respondents experiencing relapses in symptoms triggered by physical or mental activity, or stress.^15 16^ Health challenges in Long COVID, such as PESE overlap with other post-viral illnesses such as myalgic encephalomyelitis / chronic fatigue syndrome (ME/CFS) characterized by persistent and sometimes fluctuating health challenges triggered by physical or cognitive exertion.^17-19^ Collectively, these health challenges are referred to as ‘disability’, broadly defined as any physical, cognitive, mental and emotional symptoms and impairments, activity limitation, and challenges to social participation.^20 21^

While evidence from similar and overlapping post-viral illnesses like ME/CFS indicate persistent and potentially life-long disability,^22 23^ the long-term trajectory of Long COVID remains unknown. Hence, conceptualising disability in Long COVID is essential for better understanding the lived experiences and health-related challenges of people living with and affected by Long COVID to inform effective healthcare and rehabilitation approaches and interventions to enhance clinical practice, policy, and research. Our aim was to describe the experiences of disability among adults living with Long COVID in Canada, Ireland, United Kingdom, and United States.

## METHODS

### Study Design

We conducted a qualitative descriptive study involving online semi-structured interviews. This study was approved by the University of Toronto Health Sciences Research Ethics Board (REB) (Protocol #41749). This study is part of a larger study to establish a conceptual framework of episodic disability and patient-reported questionnaire of disability among adults living with Long COVID. More details on the study protocol are published elsewhere.^24^

### Patient and Public Involvement

This study is a community-clinical-academic collaboration among the Long COVID community, clinicians, and researchers in the field of Long COVID, rehabilitation and episodic disability.^25^ This study is a collaboration with Long COVID community networks and organizations including: Long COVID Physio (DAB, CT),^26^ Patient-Led Research Collaborative (HW, LMcC),^27^ COVID Long-Haulers Support Group Canada (SG),^28^ Long Covid Support (MOH),^29^ and Long COVID Ireland (NR, RS)^30^ represented by persons with experiences living with Long COVID, who were involved in all stages of the research design, sampling strategy, recruitment, analysis and interpretation of study findings. The Core Long COVID Community Collaborator Team met monthly to discuss sampling strategy, recruitment process, and characteristics to inform purposive recruitment.

### Participants

We included adults who self-identified as living with Long COVID, defined as signs and symptoms that develop during or following an infection consistent with COVID-19 which continue for 12 weeks or more and are not explained by an alternative diagnosis.^3^ We used targeted recruitment via community organizations and networks noted above to recruit 10 participants per country. We used purposive sampling in order to recruit a sample with diversity in age, gender, race/ethnicity, sexual orientation, and duration since initial presentation of COVID-19.

### Data Collection

We conducted online semi-structured interviews using Zoom in Canada, United States (KKO, BT, KMcD), Ireland and United Kingdom (NSS). Using a semi-structured interview guide, the principal investigator (KKO) and research coordinators (BT, KMcD, NSS), all of who were female and physiotherapists, asked participants about their experiences living with Long COVID, and the way in which they experienced their health-related challenges (Supplemental File 1). At the end of the interview, we asked participants to illustrate their trajectory of health experiences living with Long COVID over time.^31-33^ Following the interview, we administered a demographic questionnaire with a link via Qualtrics,^34^ an encrypted online questionnaire platform in order to describe personal, health and COVID-related characteristics of the sample. All interviews were audio recorded and transcribed verbatim. We used the visual illustrations to facilitate participants’ description of their disability experiences during the interview and supplement our understanding of the interview data for analysis.^32^ Using drawings or visual representations can foster discussion and enhance participants’ elicitation of their perceptions and experiences of illness.^33^

### Consent

All participants provided informed written verbal consent. Participants were offered a $30 CAD / $20 USD / €20 / £15 electronic gift card for their participation in the study. The interviewer followed up with each participant the day following the interview to check in on the process and to provide suggestions for support services if needed.

### Analysis

We conducted a group-based qualitative analysis using content analytical techniques.^35^ All transcripts were coded line-by-line using a coding framework informed by the Episodic Disability Framework,^20 36^ while allowing for additional codes to emerge from the data. A sub-set of 10 transcripts were independently coded by a second reviewer (KMcD). Five team members (KKO, DAB, SCC, NSSC, PS) reviewed a sub-set of four transcripts and provided higher level categories, impressions and reflections from which to contextualize the data. The community members of the team met on two occasions to review the coding summary and graphical illustrations, review and discuss participant summaries (n=15), discuss preliminary findings related to episodic nature and uncertainty, and provide reflections of validation and interpretations of the findings. We discussed the participant drawings illustrations in conjunction with their corresponding interview data to better understand their descriptions of their trajectories of episodic disability over time.^32 33^ We used NVivo software to facilitate data management.

## RESULTS

Forty adults living with Long COVID (10 per country) participated in an interview between December 2021 and May 2022. Interviews were approximately one hour in duration; five participants split the interview into two separate sessions; 34 participants used visual illustration to describe their experiences with Long COVID. The majority of participants were women (63%), white (73%), heterosexual (75%), with the majority (83%) living with Long COVID for more than 1 year, and most (93%) experienced at least one relapse in their symptoms.

**Table 1.**
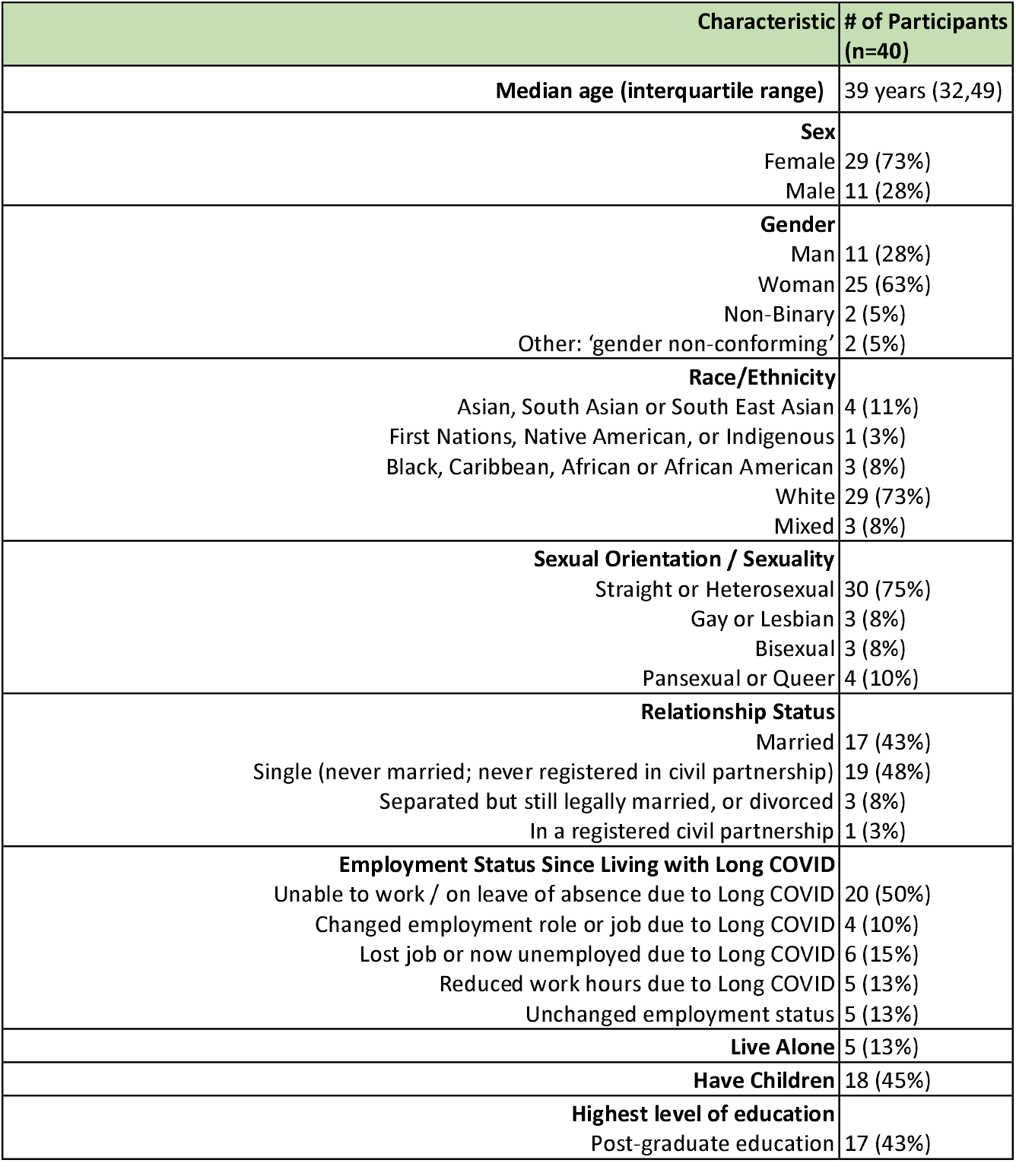
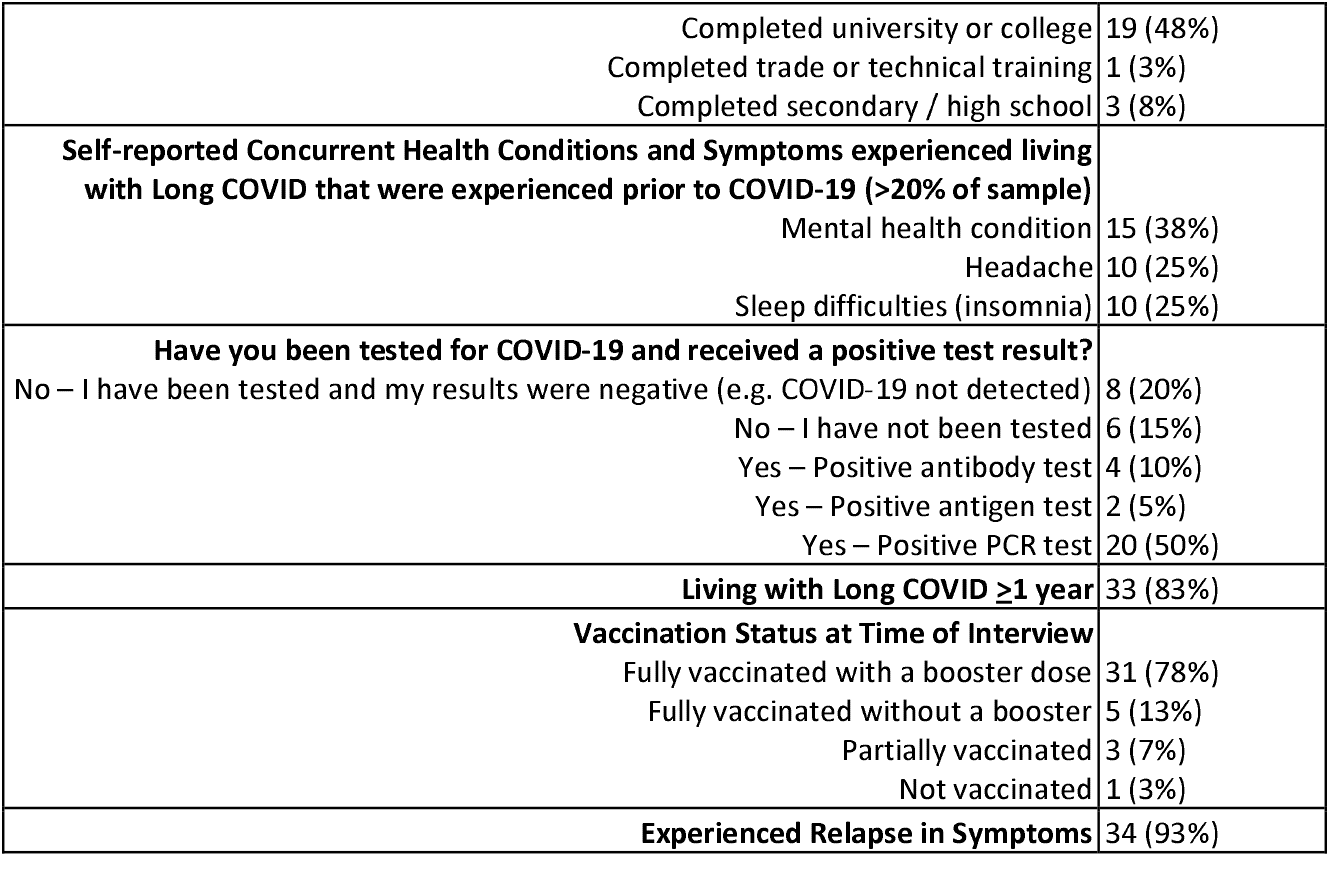
Characteristics of Participants (n=40)

Participants described their experiences of disability as episodic characterized by a diverse range of health-related challenges, resulting in short-term and long-term fluctuations in health, some of which may be unpredictable in nature, impacting their ability to plan for the future. Episodes represented marked periods in the trajectory of illness with worsening symptoms, and deteriorating health including physical and cognitive health challenges, difficulties with daily living, and difficulty engaging with work, family and social life. We specifically describe the episodic nature of disability and its uncertainty, with supportive quotes, below.

### Episodic Nature of Disability Living with Long COVID

#### A Episodic Terminology

##### A.1. Terminology

Participants described their Long COVID trajectories as episodic, referencing fluctuations in symptoms and severity over time. Episodic disability with Long COVID was characterized as living with ‘ups and downs’, ‘flare-ups’, ‘peaks’ followed by ‘crashes’, ‘troughs’, and ‘valleys’, likened to a ‘yo-yo’ ‘rolling hills’, ‘spiral’, and ‘rollercoaster ride’ with ‘relapsing / remitting’, ‘waxing and waning’, and ‘good days and bad days’, representing ‘fluctuations’ in health (Figure 1).

**Figure 1.**
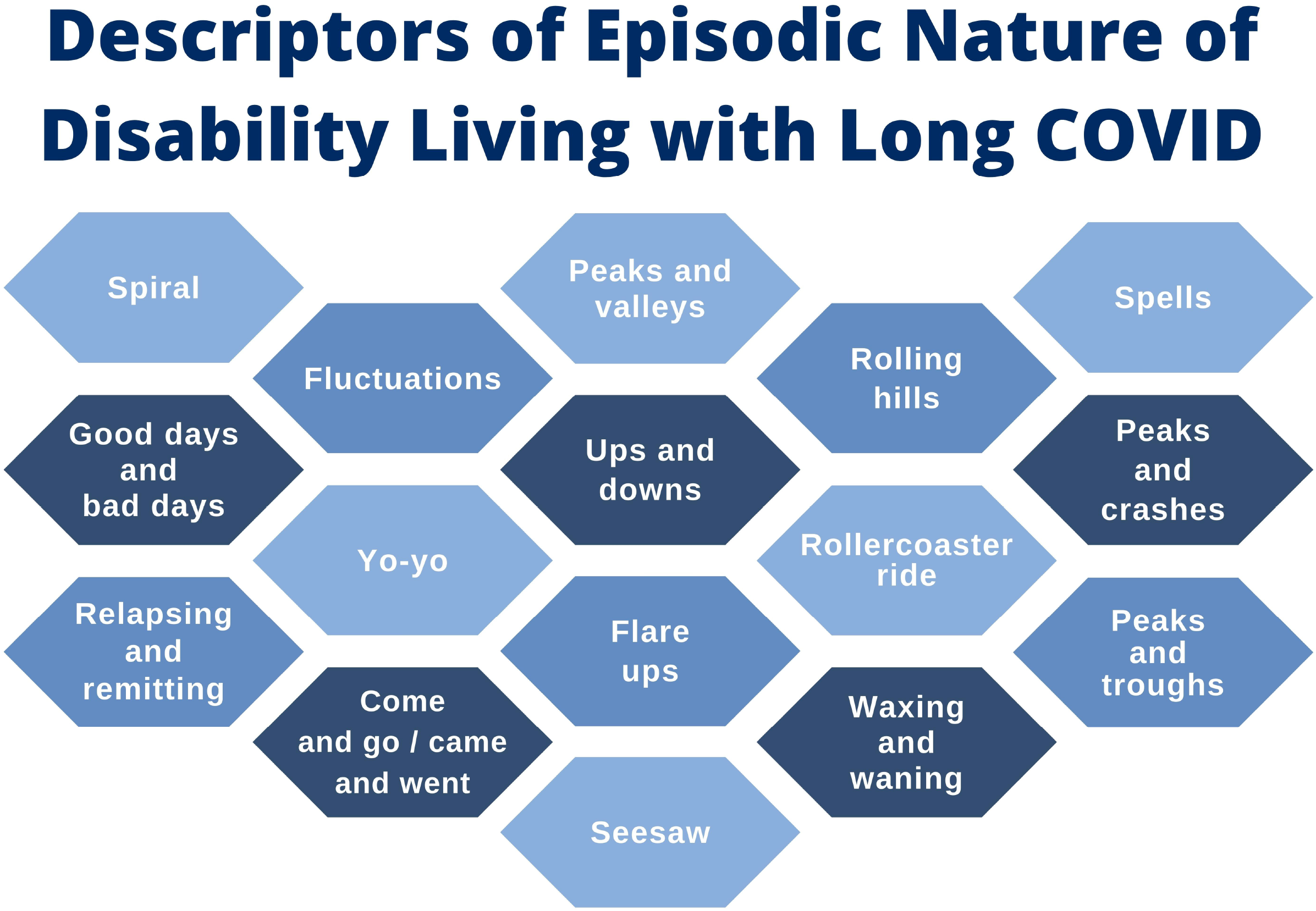
Descriptors of Episodic Nature of Disability Living with Long COVID

> *“I liken it to a rollercoaster ride and you’re like up and down… there are good days and bad days …. It’s kind of like rolling hills …. some days are good, some days are bad but the intensity of the pain and the symptoms aren’t as intense”*. [P24]

All participants described the episodic nature of illness, some resonating closely with the term as this participant stated: “*I have relapses all the time… the name like episodic disability is actually the best description that I’ve heard of it because that’s exactly how it feels”* [P27]

Choice of terminology used by participants to describe their episodic disability reflected the amplitude of an episode, described as a big picture episode (e.g. relapse, crash) considered more severe with greater disability, compared with a less severe daily fluctuation (e.g. good day and bad day, waxing or waning). Many described the episodic nature of disability as a series of crashes, followed by periods of improved health:

> *“It’s episodic in the way that your symptoms come and go depending on what you’re doing… I think it’s episodic and then you get feeling a bit better, you try a bit more and you crash, you get feeling a bit better, you try a bit more and then you crash… think that I see as episodic because I’ve been up and down bouncing around like a yoyo for two years…I agree that it’s episodic. I could be feeling fine physically and doing okay and then something really stressful happens… Then I’m going to suffer the next week.”* [P47]

##### A.2. Visibility and Invisibility of Episodic Disability

Some participants discussed the challenges of ‘invisible’ features of disability whereby friends, family and employers struggled to understand the episodic nature of their health challenges. Fatigue, headaches, cognitive dysfunction, and emotional health challenges were not visible or constant, which sometimes made it difficult for participants to articulate and have their health challenges recognized as a disability.

> *“[I]t’s very difficult living with this invisible disability because people look at you and say ‘oh well you look great’. And it’s like yeah, but I’m all screwed up inside. I can’t walk around the block without getting chest pain I haven’t picked up a vacuum cleaner for two years*.*”* [P47]

> *“You look perfectly fine but you desperately need a seat. Like people aren’t going to stand up for you just generally right. Like they’ll just assume you’ll be fine*.*”* [P66]

#### B Episodic Disability as a Continuum

Participants described episodic disability spanning over the long term (since the acquisition of acute COVID-19), as well as fluctuating on a weekly or daily basis, or within the course of a day. Participants described health challenges (disability dimensions) as “clusters”, experienced concurrently, with differing presence, severity, and duration of episodes.

##### B.1. New and Concurrent Exacerbated and Persistent Disability

Participants described disability as a new onset of health challenges since COVID-19; health challenges that were pre-existing (prior to) COVID-19; and those exacerbated since COVID-19. For individuals living with pre-existing disability (either permanent or episodic in nature), their health challenges were now compounded with those associated with Long COVID, furthering the complexity of their disability. Of note, not all health related challenges were experienced as episodic, some were more persistent, consistent or stable over time. For instance, fatigue was commonly expressed as a consistent health challenge with episodic severity: “*The fatigue is constant but it comes in waves…I’m always tired but some days I wake up and I’m just absolutely exhausted*.” [P15]

Another participant illustrated their relapsing disability with Long COVID over two years characterized as “clusters” and “spikes” of disability, while pain was persistent (orange line) within the broader episodic nature of disability as described in this quote and visually described in Figure 2:

**Figure 2.**
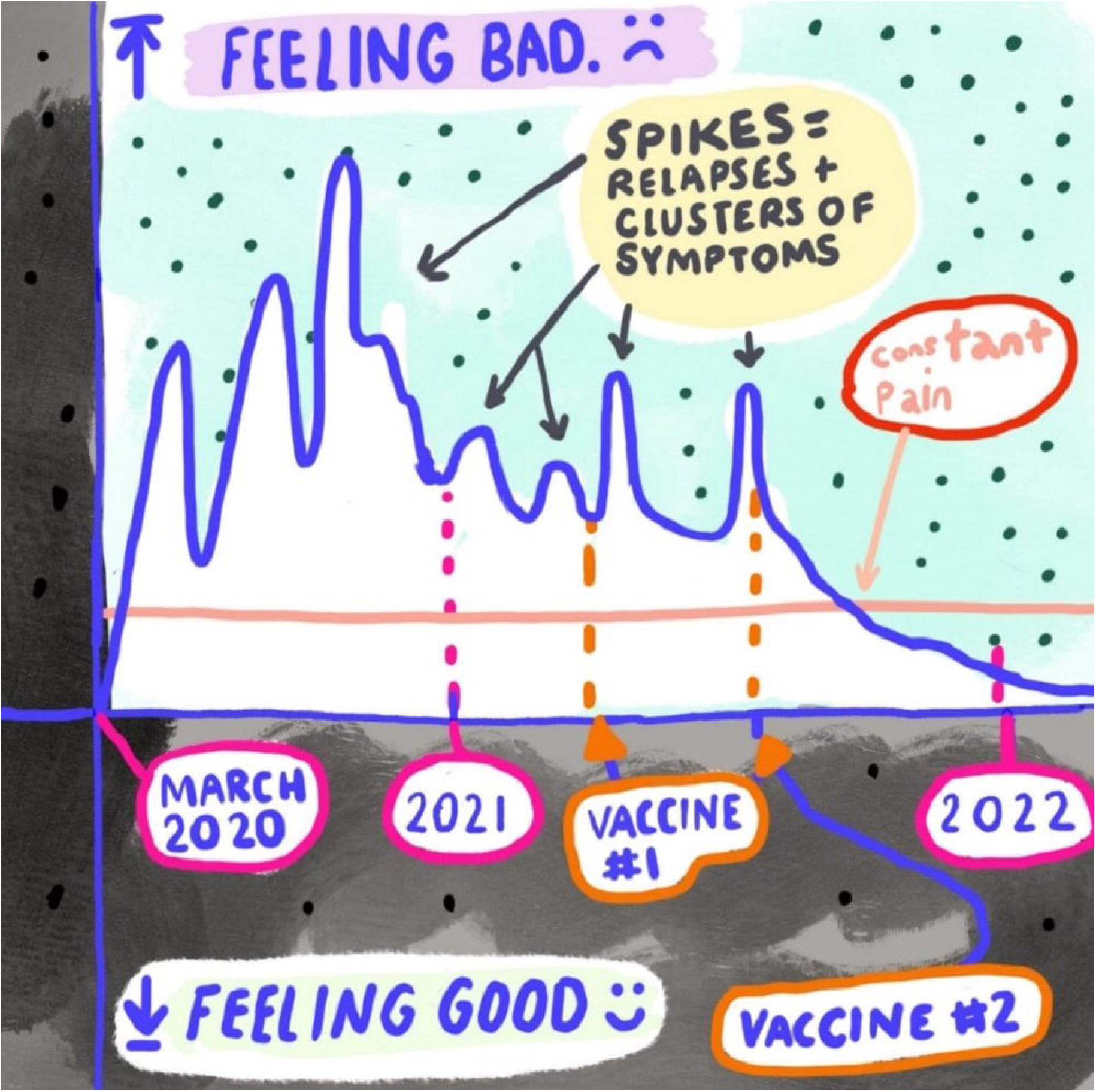
Visual illustration of experiences living with Long COVID over years – episodic disability (P3)

> *“From the beginning what became clear was that the relapsing nature of it, up until the vaccine, that relapsing never stopped… In June I had a huge reaction when I was bed-bound… that relapse was worse than ever before because I was never bed-bound when I first got really sick. In June/July, that relapse was… the worst that I’ve ever been… I’ve been really lucky that since my second vaccine… I’ve noticed that I’m not relapsing with my energy*.*”* [P3]

##### B.2. Multi-dimensional Nature of Disability

Participants described the multi-dimensional nature of episodic disability, comprised of physical, cognitive, mental and emotional health challenges, resulting in difficulties with daily function, and social participation. This participant described how the episodic nature of disability differed depending on the type of health challenge (or dimension of disability), with some more stable compared with others which fluctuated in greater severity and frequency over time each articulated with a different colour in the graph (Figure 3).

**Figure 3.**
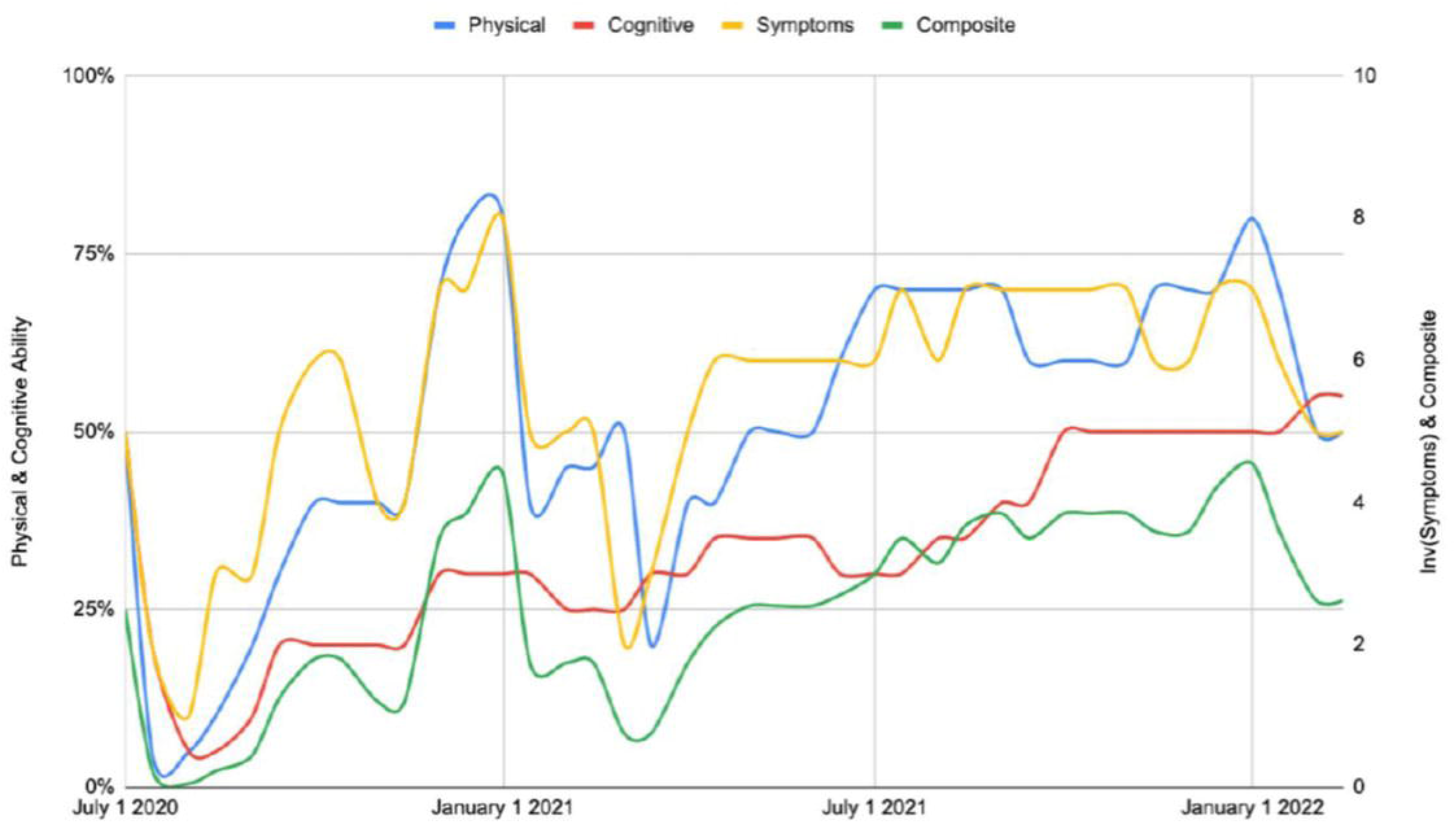
Visual illustration of experiences living with Long COVID - multidimensional nature of episodic disability (P26)

##### B.3. Daily Fluctuations in Episodic Disability

Participants described the good days and bad days living with Long COVID, the ups and down, and rolling hills that fluctuated on a daily basis (‘good days and bad days’) as well as within the course of a day. As this participant states:

> *“I liken it to a rollercoaster ride and you’re like up and down… there are good days and bad days*.*… my symptoms are still there but it kind of goes down this way right. So it does this now I think and the ups and downs aren’t so pronounced. It’s kind of like rolling hills if you will …So some days are good, some days are bad*.*”* [P24]

This participant illustrated the trajectory of their illness over the course of the day, its unpredictability and the strategies they implemented to mitigate symptoms (Figure 4):

**Figure 4.**
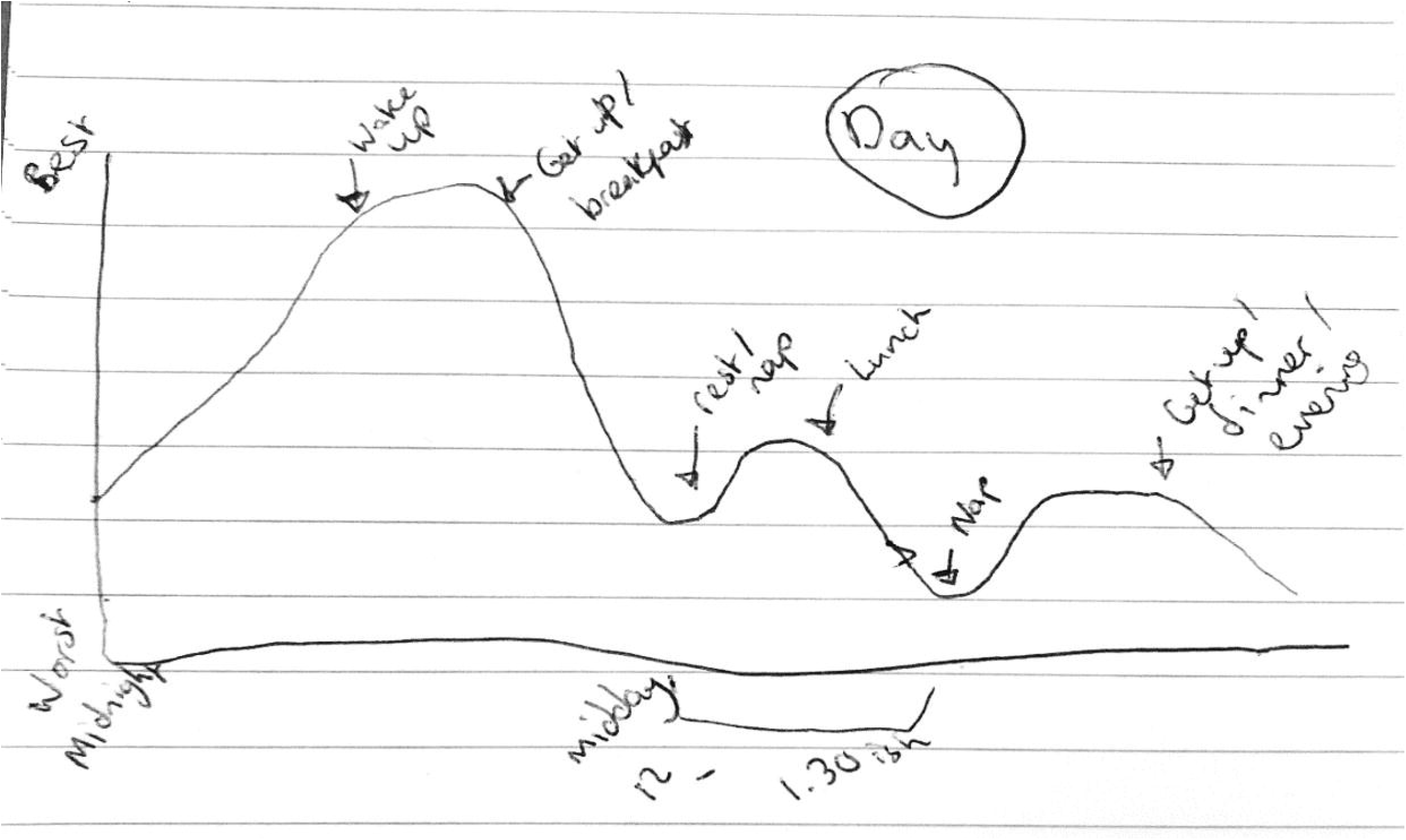
Visual illustration of experiences living with Long COVID – daily episodic nature of disability (P48)

> *“I might struggle to get out of bed in the morning. Waking up is difficult. I would take quite a while to kind of sit up out of the bed… I’m not always up to have the kids get ready for school. Then I might manage to kind of putter around and do things within the home maybe until around lunchtime and then I’d start to feel exhausted again. So I’d be maybe lying down on the sofa or in the bed before I have to go and get the children from school … and then I have to do the dinner and the homework and stuff. So really I’m conserving energy a lot of the time*… *I can’t really tell on a day to day basis what that’s going to be like, whether I’m going to be able to manage that or not”* [P48]

#### C Changes in Episodic Disability over Time

##### C.1. Persistent or Stable Disability

Some participants described permanency of health challenges, stating they were living in a constant state of disability, reflected by the loss of function compared with their baseline level of health, stating “it’s still always there, it’s just the severity of it changes” [P65]. They described how stable features of disability may co-exist with episodic disability with Long COVID:

> *“I consider myself disabled 24/7. I don’t think I have points where I have the abilities that I previously had…*.. *So I don’t really view my disability as episodic in that way. I’m always at this minimal state and sometimes I’ve got to make that state even more minimal but it’s not like I go from ‘oh today I can do all these things’. No, there’s no day like that*.*”* [P22]

##### C.2. Episodic Disability Trends of Improvement or Deterioration

Some participants described an overall improving trend in health over time living with Long COVID, super-imposed by periodic episodes, flare-ups or daily fluctuations in disability over time.

> *“Currently the challenges are more episodic in that if I have daily work activities, exercise or a certain volume of activity that I used to be able to perform regularly, I tend to have some sort of flare-up …*.*So those symptoms came and went repeatedly. The general trend is it’s a little bit better now but it’s still limiting in that I have to be very careful about how much activity I do in a day or in a week”*. [P2]

Nevertheless, improvement in disability often came with the trade off of pacing and having to scale back on energy expending activities previously tolerated prior to Long COVID, such as employment or engagement in social activities: *“I’m very lucky to see an incredibly slow upward trajectory. I think that is partially due to doing less”* [P26].

##### C.3. Navigating Episodes of Disability

Participants described a reserve of energy that they had on a given day, and if they went beyond that reserve, it would trigger an episode. Energy, and consequently functional ability, was a finite resource. Participants described the effort required to continually balance their level of activity with energy levels on a given day (which may fluctuate) and the careful balance and navigation required to ensure they did not relapse or crash with more disability.

> *“I’m hoping that I’ll just keep slowly crawling upwards… I could be feeling fine physically and doing okay and then something really stressful happens then I’m going to suffer the next week… If I laid around all the time and didn’t challenge myself I probably wouldn’t have as many symptoms … But then I’d get way deconditioned. I think I’ve just learned how to hopefully make those peaks and valleys not as far apart and to have a bit more stability”*. [P47]

Others described episodic disability as a “moving target” as they try to navigate strategies to mitigate or prevent episodes. This came with underlying uncertainty of whether improvements in health were attributed to recovery from Long COVID or refining their strategies to mitigate episodes of disability.

> *I feel like it’s a moving target. I’m still trying to figure out two years later what I’m capable of, although I am getting better or I feel like I am. But sometimes I feel like I plateau and how much is also getting better versus just better at pacing*. [P38]

Overall, episodic disability trajectories reflected with an absence of a progressive path from illness to wellness recovering from COVID-19. Most participants exemplified fluctuations in health that despite having good days, they had not returned to a baseline level state of health, highlighting the complexity of the experiences living with Long COVID.

In summary, participants experienced episodic disability, not as an all or nothing concept of complete wellness and complete illness, but a continual state of health challenges with changing presence, severity and duration of episodes over time.

### Uncertainty Living with Long COVID

The episodic nature of living with Long COVID was interconnected with the concept of uncertainty. Participants described the sometimes unpredictability of fluctuating health challenges, worrying about the future, lack of knowledge on the causes of and treatments for Long COVID, and the impact that uncertainty of Long COVID had on their overall health and future life decisions. Participants described concerns of not knowing what to expect, as they experienced new and relapsing clusters of symptoms, and not knowing the nature or course of their illness over the short or the long term:

> *“The biggest challenge is not knowing when they’re [episodes] going to happen, how long they’re going to last or how much I can get away with before causing it, if I am truly the cause of one of these episodes*. [P2]

Another participant articulated the uncertainty of episodes, by referring to them as *“my spells because it feels kind of like a mystery of what’s going on”* [P38]

Uncertainty of disability living with Long COVID occurred in the form of day-to-day uncertainties (will I be able to get out of bed today? will I be able to keep my plans?), as well as worrying about the future in relation to the ability to work, have a family, perform family and social roles, activities (will I be able to go back to work? will I ever return to my ‘old self’? will I be able to care for my children?). As one participant described, the daily uncertainty: *“you never really, know when it’s going to be a good day and when it’s going to be a bad day”* [P64].

Despite efforts to predict when an episode may arise, participants referred to underlying and persistent uncertainty of their episodic trajectory living with Long COVID and the impact on their ability to engage in social participation:

> *“I couldn’t really commit to even social plans. there were some days where I would have to cancel last minute because it just wasn’t a good day there’s just not a lot of predictability despite me trying to track everything and log all of the triggers and symptoms and durations, it’s still hard to predict*.*”* [P2]

The persistent unpredictability of Long COVID had implications on employment, financial security, and broader implications on future health and life decisions, such as family planning or returning to work:

> *“If I knew that I could get away with two days of part time work maybe not back to back…. and that wouldn’t flare up my symptoms, I would probably go back to doing that if I could. But it’s so inconsistent and it’s so hard to predict in that sense that I can’t really commit to anything employment-wise. …There’s just not a lot of predictability*.*”* [P2]

This participant described their uncertainty of episodic disability in relation to ability and readiness to return to work:

> *“How will you know you’re ready to return to work? [I’ve been told that] you have to be able to get through the basics of your own day to day life and be fully functional at it with capacity to spare before you can try and put your toe back into the working world. I haven’t gotten there in the 14 months since my initial infection and it’s really hard to know if I ever will. Maybe if treatments come along that can put in check those … crashes that I experience … Maybe we can find what the underlying cause is that keeps me sick and keeps me relapsing. Then maybe it will be conceivable. But as is I do not experience remittance for nearly long enough for that to be feasible…*.*It’s just I’m not in a stable enough place for that to be a realistic goal*.*”* [P40]

This participant described the consequences of uncertainty related to causes of their episodes, their ability to engage in daily functional activities, and social activities such as travel or leisure:

> *“Fluctuations, it’s unpredictable really… last year I was going through a period where I thought I was improving and I thought I was on like an upward trajectory and I was I suppose until a certain point. So I got to a stage where I was able to go walking, go on walks for like up to an hour without crashing…*.. *And then it got to a point where the walk, going for a walk, was making me crash the next day… I’ve kind of just been down. I don’t know if … I’m stable or going in towards a downwards trajectory. It’s hard to tell really… I really don’t know what caused that shift from like improving to losing that*. [P64]

Uncertainty was also closely linked to fear of not knowing what the nature and severity of an episode might be, the potential for new onset of symptoms, its consequences, and the widespread fear of not recovering from Long COVID:

> *“At first I had no idea what was happening and most people didn’t and it was terrifying to see these new symptoms …and just have zero explanation for what was happening. I also experienced a lot of fear and uncertainty around how long is this going to last…it’s a very frightening prospect to think of that number never getting better*.*”* [P26]

### Furthermore, participants expressed uncertainty of the underlying causes of Long COVID

*If you can understand at least the basics of why (Long COVID happens)… the underlying mechanism of whatever is happening with you, then you can accept it and deal with it and then try and you know build your life to kind of pace yourself around it. But when you have no idea… you can’t*.*”* [P1]

Uncertainty of Long COVID also was a concept participants stated were experienced by health care providers, insurers and employers as they also grappled with the unknown of the underlying cause, treatment, and broader trajectory of Long COVID. This participant refers to worrying about their long-term health while recognizing parallel uncertainty faced by health providers:

> *“The more months that go on like it’s… as I’m getting the nearly 12 months now I am getting to get a bit worried like will I get back to where I was. I’d take 90%. If you offered it to me there, I’d take it. My doctor even said that to me. He said like there’s no guarantee I’ll get back to… I hope I can get back to 90%. I’d take that*.*”* [P14]

Participants referred to their experiences of uncertainty when working with health care providers, and undergoing diagnostic testing and results coming back normal despite ongoing persistence of disability that further contributed to uncertainty and worrying about the future:

> *“if you go get a test you’re like ‘I hope nothing’s wrong’. I’ve gotten to the point where I’m like ‘I hope something is wrong’ because over the last two years I’ve had too many tests and it’s like ‘oh everything is normal, your heart is good’. … okay well I’m having all these symptoms which fit everything or whatever but normal test results which is supposed to be a good thing but over a couple of years of that and still feeling like I do… It’s depressing… at least I want something that’s treatable. Like with the MRI they said ‘oh no, it’s arthritis in your lower back’. I’m in a lot of pain but that’s treatable. Instead of getting a test, ‘oh everything is fine’ and you’re just in a lot of pain. That’s a problem*.*”* [P45]

Participants distinguished between aspects of disability that were more controllable than others and the strategies they employed to create greater predictability in their lives, to prevent or anticipate episodes of disability in the context of some of the less controllable episodes of disability.

> *“On a day to day basis…I know that if I take that walk to the clinic, then that is the only thing I can do that day. So yes, I can pace my activities on a day-to-day basis. The thing that is unpredictable is the underlying fluctuations which happen on a weekly and monthly basis. So if you’re in a trough period then you know you have no options apart from just trying to recover from the crash. So it’s literally lying in bed. There doesn’t seem to be yet a way of predicting when that might happen in my case*.*”* [P15]

Participants described drawing from their experiences living with Long COVID as a strategy to gather information on the ‘certainty’ that came from understanding the trajectory of a given ‘crash’ or episode of disability. For those living with Long COVID longer, some had a good understanding of the predictability of some episodes of which they could establish strategies to mitigate or prevent in the future:

> *“If I do feel bad, it’s unpredictable*…*the time I would be feeling low just comes out of nowhere. But that probably is linked to maybe doing a bit too much activity and then I would say ‘okay I know I can’t do that just yet’, … and draw it back to my little baseline of doing basically nothing, and then I’m fine again*.*”* [P1]

## DISCUSSION

This work articulates the complexity of episodic disability experienced among people living with Long COVID. All participants described health challenges experienced with fluctuating severity and duration over time. Participants characterized episodic disability as ‘clusters of symptoms’ that can overlap and ‘fluctuate and change over time’, described as ‘prolonged’, ‘relapsing and remitting’, that may fluctuate within the day, between weeks, or over a longer continuum of months to years. Results validate our earlier work that characterized Long COVID as an episodic health condition^37^ and the importance of the inclusion of ‘relapsing and remitting’ in the World Health Organization definition of Long COVID.^3^ The concept of episodic disability was originally derived from the context of HIV, and applicable to other health conditions where challenges can fluctuate daily or over longer periods of time.^20 36 38^ The Episodic Disability Framework describes disability as multi-dimensional, and episodic in nature.^20 36^ Utilising an existing framework of episodic disability developed from, and validated with others living with episodic illness, provides a foundation for understanding disability experienced among people living with Long COVID. This offers a novel way to conceptualise disability experienced by people living with Long COVID.^39 40 37 41^

Episodic disability among participants was conceptualized as a continual state of change, or fluctuations along a continuum of varying severity, duration, type and presence of health related challenges in between the extremes of health and illness. The episodic nature of disability has been documented in examinations of physical and cognitive health challenges after COVID-19,^42^ and authors acknowledge the fluctuating nature of symptom flare ups (bad days) followed by controlled periods of less symptoms (good days), that may vary (or fluctuate) over time within the same individual.^43^ The potential ‘invisible’ feature of episodic disability can further complicate the ability to articulate health challenges experienced by adults living with Long COVID to family, friends, employers or health providers.^44^ This aligns with recent recognition of Long COVID as a disabling condition,^45^ and the critical need to extend social supports and policies grounded within disablement framework, informed by people who live with disabilities as a basis for progressive change.^44^

The term ‘episodic disability’ was perceived as way to legitimize living with the health challenges of Long COVID while recognizing that disability is not experienced as an all (disabled) or nothing (non-disabled) concept. Episodic disability may be synonymous to dynamic disability, a term used to describe progressive, recurrent or fluctuating limitations.^46^ This may translate to an individual possessing intermittent ability for work capacity or ability to manage sustained or extended periods of workload due to their fluctuating health condition.^47^ Many persons living with disability experience their disability as dynamic, meaning progressive, recurrent or fluctuating over time,^46^ which can increase with age, and more commonly experienced among women compared with men, and have implications on employment.^48^ However, an important finding from this study, is that not all health challenges were experienced as episodic, some were more stable or persistent, with fluctuating severity and trends of improvement or deterioration over time highlighting the importance of recognizing not all health challenges in Long COVID are experienced as episodic or of equal severity and duration. Given the importance of terminology to provide clarity of understanding and communication among community and health providers, we recommend the use of the term ‘episodic disability’ to collectively characterize the multidimensional and potentially episodic and unpredictable health-related challenges associated with Long COVID.

Perceptions of disability experienced in Long COVID may also change over time. Tran and colleagues (2020) documented fluctuating presence and burden of symptoms with Long COVID over time, revealing a timeframe whereby initially, the burden of disease decreased as several symptoms disappeared, followed by an increased prevalence of burden of symptoms, suggesting correspondence to the onset of chronic illness.^49^ Trajectories of episodic disability have been explored among adults living with HIV, highlighting distinct dimensions that may be experienced as increasing, decreasing, stable and fluctuating over time.^50^ Future longitudinal studies are needed to measure disability over time in order to characterize its nature, severity and episodic nature over time with Long COVID and to explore the burden of living with sustained episodic disability and uncertainty over time and strategies to mitigate or prevent disability.

Some of the data describes the types of physical, cognitive, and social forms of disability that were experienced by participants. In this paper we focused on the episodic and uncertain nature of disability and its trajectory. The exploration of the dimensions (or types) of health related challenges that comprise disability, the contextual factors that exacerbate or alleviate disability, and triggers of disability is a focus of future work.^24^

Uncertainty and worry about the future was a key feature of disability experienced by participants, described as the inability to predict episodes, the overall trajectory of their illness, and impact on future life decisions. With Long COVID, the impact of living with the uncertainty of when an episode of more severe symptoms might arise, the severity and duration of that episode, and the long-term implications on health and life decisions, including the financial, practical, and emotional consequences are unknown. Uncertainty with Long COVID can span multiple facets, including as a dimension of disability (uncertainty when an episode might arise, its severity and long term implications on health), diagnostic uncertainty (diagnostic clarity among those without a formal PCR or diagnostic test), financial uncertainty (if when and how able to return to employment), uncertainty among health care providers (how to assess and treat, or epistemic injustice), and among employers and insurers (how to accommodate).^44^ Living with uncertainty was a key theme emerging from a qualitative study, highlighting fear of whether recovery was possible among adults with Long COVID.^51^ Uncertainty further may persist as people with Long COVID experience uncertainty about what might occur in the event of COVID-19 re-infection. Uncertainty is a key dimension of disability and predictor of mental and emotional health and social inclusion in the context of HIV ^52^ and features of uncertainty were similarly documented with older adults aging with HIV, pertaining to the source of health challenges, episodic nature, uncertainty of health care providers knowledge and skills, and financial uncertainty.^53^ The lessons learned from HIV can offer insights into uncertainty experienced during the new onset of a pandemic.^41^

While uncertainty and concerns about the future living with Long COVID was a prominent theme in this study, having uncertainty doesn’t make an individual disabled, nor does it suggest that episodic disability with Long COVID originates from one’s mental state of health. Of note, uncertainty may not always be negative; evidence highlighted the role in embracing uncertainty as a positive, and understanding a person’s episodic trajectory may help to tailor strategies or interventions that may mitigate episodic dimensions of disability.^54 55^ Similarly, pre-existing health conditions and symptoms reported by participants living with prior to COVID-19 should not be interpreted as associated with or predictor of Long COVID, rather they provide context for understanding the comorbidities that may be experienced among the sample in addition to the disability experienced with Long COVID. As evidence continues to emerge on the long-term outcomes and trajectory of Long COVID, and the underlying mechanistic causes of symptoms and disability,^43^ uncertainty with Long COVID will change. Clarity of diagnostic criteria can help to reduce uncertainty and validate experiences of persons living with Long COVID including those without formal testing at the time of COVID-19 infection.^56^ In the meantime, uncertainty should be recognized as an integral component of disability experienced among individuals living with Long COVID so that healthcare providers can better understand and support those living with uncertainty in the context of chronic illness. This should be considered in the context of concurrent uncertainty experienced in the broader environment among caregivers, health providers, employers and insurers in the evolving field of Long COVID.

### Implications for Practice, Research and Policy

Having clear recognition and diagnostic criteria of episodic disability associated with Long COVID will help to recognize the health challenges experienced with Long COVID to access timely clinical assessments and treatments, safe rehabilitation services, models of care, disability justice, employment rights, and income support. Knowledge on Long COVID and episodic disability in health professional educational curriculum, and education of current and future health providers employers, insurers, and policy stakeholders, are needed to equip stakeholders with a better understanding of the experiences to inform equitable, and patient-centred approaches to models of service delivery for people living with Long COVID.^57^ The episodic nature of disability highlights the role for rehabilitation to help prevent, address, and mitigate disability, and to enhance health outcomes in Long COVID. Rehabilitation should be disability focused, safe, informed by existing post-viral illness research, goal-oriented, person-centred, focused on function, tailored to an individual’s goals, abilities, and interests.^58 59^ Future measurement of disability is important to determine what dimensions of disability may or may not change over time.

Disability is impacting the health of individuals living with Long COVID in addition to the implications on society as a whole.^60^ There is a critical need to understand, measure and recognize the impact of disability among people living with Long COVID.^61^ Development of a robust research agenda on disability and rehabilitation involving citizen scientists is vital for establishing evidence for safe and effective rehabilitation models of care in Long COVID.^62^ Community-engaged and disability-inclusive approaches to addressing Long COVID episodic disability will help to foster timely and relevant interventions and approaches to healthcare, rehabilitation, and the application of research evidence about appropriate service planning and delivery.^63^ Opportunities exist for partnering with related chronic and episodic conditions like Myalgic Encephalomyelitis/ Chronic Fatigue Syndrome (ME/CFS), Lyme disease, and dysautonomia to advance rehabilitation, employment and insurance policy and programming related to Long COVID rehabilitation.^64^

### Strengths and Limitations

Strengths of our study included our community-engaged and international approach, involving persons living with Long COVID across five community networks in four countries who were involved in all aspects of the study. This work reflects an academic-community-clinical collaboration embedded in a strong foundational knowledge and lessons learned from the field of episodic disability and rehabilitation.^41^ Our large sample size of 40 interviews and use of graphical illustrations complemented the interview data to comprehensively describe the experiences living with Long COVID. By recruiting through collaborator organizations by members of the team living with Long COVID, we achieved equal representation across the four countries in the sample. We analyzed the data as a collective sample as our aim was not to compare or contrast by country. The majority of the participants were women, white, and living with Long COVID for more than a year, who identified as living with Long COVID and its associated disability, and were able to participate in an online interview, hence participants may not reflect the broader population of adults living with Long COVID.

## CONCLUSIONS

In summary, experiences of disability were described as episodic, characterized by fluctuating health challenges, which may be unpredictable in nature highlighting intersections of uncertainty among this sample of adults living with Long COVID. Results provide a better understanding of the lived experiences of disability among adults living with Long COVID that may help to inform approaches for future healthcare and rehabilitation.

## Data Availability

All data produced in the present work are contained in the manuscript.

## ACKNOWLEDGEMENTS

We thank the participants for their contributions to this study and the community organizations who collaborated in this work. We thank Laura Bassi, FisioCamera, for their role in developing the Figure 1 graphic.

## FUNDING

This study was supported by the Canadian Institutes of Health Research (CIHR), Emerging COVID-19 Research Gaps and Priorities Funding Opportunity (Funding Research Number #: GA4-177753), 160 Elgin Street, Ottawa, Ontario, Canada, K1A 0W9). The CIHR had no role in the design of this study and will not have a role during its execution, analyses, interpretation of the data, or decision to submit results. Kelly K. O’Brien is supported by a Tier 2 Canada Research Chair in Episodic Disability and Rehabilitation and Angela M. Cheung is supported by a Tier 1 Canada Research Chair in Musculoskeletal and Postmenopausal Health from the Canada Research Chairs Program.

## COMPETING INTERESTS

The authors have no competing interests to declare.

## ETHICS APPROVAL

This study received approval from the University of Toronto Health Sciences Research Ethics Board (Protocol Reference #41749).

## SUPPLEMENTAL FILES

Supplemental file 1 – Interview Guide - Long COVID Episodic Disability Study

## Notes

### Competing Interest Statement

The authors have declared no competing interest.

### Author Declarations

The University of Toronto Health Sciences Research Ethics Board gave ethical approval for this work (Protocol Reference #41749).

